# Detection of Measles Virus RNA in Wastewater: Monitoring for Wild-Type and Vaccine-Derived Strains in a National Preparedness Trial

**DOI:** 10.64898/2026.04.09.26350527

**Authors:** Warish Ahmed, Metasebia Gebrewold, Rory Verhagen, Melissa Koh, Jake Gazeley, Avram Levy, Stuart Simpson, Monica Nolan

## Abstract

Wastewater surveillance (WWS) is established as a vital tool for monitoring polio and SARS-CoV-2 with potential to improve surveillance for many other infectious diseases. This study evaluated the feasibility of detecting measles virus (MeV) RNA in wastewater as part of a national WS preparedness trial in Southeast Queensland, Australia, from March to June 2025. Composite and passive sampling methods were employed in parallel at three wastewater treatment plants serving populations between 230,000 and 584,000. Nucleic acids were extracted and analyzed using RT-qPCR targeting MeV N and M genes to distinguish wild-type and vaccine strains. MeV RNA were detected in both 24-hour composite and passive samples on May 26–27, 2025 from the largest catchment of 584,000 which also included an international airport. No measles cases were reported in this city or region within 4 weeks of the WS detections. These were confirmed as vaccine-derived measles virus (MeVV) strain via specific RT-qPCR assay. Extraction recoveries varied (11.5%–70.5%), with passive sampling showing higher efficiency. This is the first report of use of passive samples for detection of MeV. These findings are consistent with other studies reporting WWS results of both MeVV genotype A and wild type genotype B and/or D. It demonstrates the potential for sensitive MeV WWS with rapid differentiation of MeVV from wild type MeV shedding, including in airport transport hubs and with different sample types. Use of WWS could strengthen measles surveillance by enabling rapid detection of MeV RNA and supporting outbreak preparedness and response. This requires optimised methods which are specific to or differentiate wild-type MeV from MeVV. Furthermore, the successful detection of MeV using passive sampling in this study highlights its potential for deployment in diverse global contexts which may include non-sewered settings.

## Introduction

Wastewater surveillance (WWS) has emerged as an important tool for monitoring certain infectious diseases, such as polio and SARS-CoV-2, and shows promise for others. While it offers a non-invasive, cost-effective, and scalable approach for population level surveillance, further evidence is needed to establish its utility across a broader range of pathogens (Farkas et al. 2024; Grassly et al. 2024).

The Coronavirus Disease 2019 (COVID-19) pandemic catalyzed global interest in WWS, demonstrating its utility in detecting viral RNA from severe acute respiratory syndrome coronavirus-2 (SARS-CoV-2) in sewage and providing early signals of community transmission, often preceding clinical case reports (Ahmed et al. 2021; Lastra et al. 2022; WHO 2023). Since then, the scope has expanded to include a diverse array of pathogens, including respiratory viruses (e.g., influenza A virus, respiratory syncytial virus), enteric viruses, and emerging threats such as monkeypox virus (MPXV) and arboviruses (Ando et al. 2023; Fanok et al. 2023; Lee et al. 2024; Mejia et al. 2024; Smith et al. 2024). This expansion reflects a growing recognition of WWS as a tool which may strengthen traditional surveillance approaches, particularly in settings with limited diagnostic capacity or delayed case reporting.

Measles virus (MeV) is a highly contagious, single-stranded RNA virus of the genus *Morbillivirus* (family *Paramyxoviridae*). It is exclusively a human pathogen, transmitted via respiratory droplets and aerosols, with a basic reproduction number (R_0_) among the highest of any known infectious agent. While R_0_ is often estimated between 12 and 18 in susceptible populations, a systematic review found that estimates vary considerably across settings (Guerra et al. 2017). Measles typically presents as an acute febrile illness with characteristic symptoms including cough, coryza, conjunctivitis, and a maculopapular rash (Alves et al. 2020). However, complications such as pneumonia, encephalitis, and death can occur, particularly in young children, malnourished individuals and immunocompromised individuals (Mina et al. 2015).

Measles remains a significant public health concern despite the availability of a safe and effective vaccine (Parums 2024). Measles is a global elimination target. The WHO Measles–Rubella Strategic Framework 2021–2030 underscores the importance of sensitive detection and differentiation of measles virus to support verification activities and prevent re-establishment of endemic transmission (Measles and rubella strategic framework 2021-2030 2020). Global efforts to eliminate measles have led to a marked reduction in genotypic diversity, with only genotypes B3 and D8 circulating since 2021 (Bankamp et al. 2024). However, progress toward elimination has stalled in recent years, with increasing cases and outbreaks reported globally. Importantly, both measles vaccination and natural infection confer lifelong immunity, underscoring the critical role of sustained immunization coverage in achieving elimination goals (Minta et al. 2024).

Recent studies suggest that circulating genotypes may accumulate mutations within T-cell epitope regions, potentially impairing recognition by vaccine-induced CD4^+^ T cells and reducing cellular immune responses (Emmelot et al. 2025). Such changes could contribute to ongoing transmission in populations with suboptimal coverage, highlighting the need for genomic surveillance alongside vaccination efforts to monitor viral transmission and progress towards elimination (Bankamp et al. 2011).

MeV RNA is shed in respiratory secretions as well as urine during acute infection (Permar et al. 2001; Gresser and Katz 1960; Sari et al. 2020). These characteristics make MeV a suitable candidate for environmental surveillance, particularly through wastewater. Measles vaccine virus (MeVV) strains belong to genotype A and MeVV RNA are occasionally detected in clinical samples following immunization, typically at low levels and for short durations (Rota et al. 1995, Washam et al. 2024). Prolonged shedding of MeVV RNA >100 days after most recent measlescontaining vaccination has been observed among 11 children in a retrospective observational study using stored diagnostic specimens (McMahon et al. 2019).

Several studies have confirmed the feasibility of detecting MeV RNA in urban wastewater samples. For example, retrospective analysis in the Netherlands first identified MeV RNA in sewage during an outbreak in 2013, with detections aligning with known transmission areas (Benschop et al. 2017). Subsequent investigations in Canada, France, Belgium, Switzerland, and the United States have applied reverse transcription digital PCR (RT-dPCR), multiplex RT-qPCR assays, and hybrid-capture sequencing to detect and differentiate wild-type MeV and MeVV strains in wastewater (Gan et al. 2025; Javornik Cregeen et al. 2025; Paulos et al. 2025; Rector et al. 2025; Roman et al. 2025; Tomalty et al. 2025). In South Africa, sentinel surveillance detected MeV RNA in wastewater of approximately half the populations districts that had no reported clinical cases, highlighting the potential of WWS to uncover silent transmission and complement traditional epidemiological data (Ndlovu et al. 2024).

In Canada, genotyping of wastewater samples confirmed the presence of MeVV RNA following immunisation campaigns, underscoring the importance of distinguishing vaccine-related signals from true outbreaks caused by wild-type MeV (Tomalty et al. 2025). Similarly, studies in USA and Switzerland demonstrated the early-warning capability of WWS, with MeV RNA detected prior to clinical case confirmation and closely tracking outbreak curves (Gan et al. 2025; Javornik Cregeen et al. 2025; Paulos et al. 2025). Collectively, these findings support the integration of MeV into WWS programs, particularly in regions with low vaccination coverage or delayed clinical reporting. In 2025, the USA and the Republic of South Africa have integrated MeV as part of their national multipathogen wastewater surveillance programs at scale (https://www.nicd.ac.za/measles-wastewater-surveillance-dashboard/; https://www.cdc.gov/nwss/rv/measles.html).

The Australian Government Department of Health and Aged Care (now Department of Health, Disability and Ageing) and the interim Australian Centre for Disease Control (iCDC) (now the Australian Centre for Disease Control (CDC)) funded a WWS pilot project, undertaken over a three-month period with collaborating laboratories and water utilities across Australia aimed to strengthen preparedness for use of WWS for emerging pathogens. The pilot study included use of multiple wastewater sampling approaches and viral pathogens to inform optimal practice. Within this framework, a targeted surveillance effort was conducted to assess the potential for detecting MeV RNA in wastewater with locations and target pathogens guided through consultation with health authorities.

This short note presents findings from the MeV WWS trial, conducted between March and June 2025 in a large subtropical city in Queensland (Qld), Australia. The objectives were to (i) evaluate the feasibility of MeV for WWS, (ii) assess the performance of sampling and laboratory methods in detecting MeV RNA, and (iii) consider the potential role of MeV WWS data within broader multimodal surveillance systems. The study also aimed to inform the design of agile surveillance responses for emerging threats inclusive of WWS and to support evidence-based decision-making in the context of national preparedness planning.

## 2. Materials and Methods

### 2.1 Wastewater Sampling

Composite wastewater samples (~200 mL) were collected over 24 hours from the influent of three WWTPs (WWTP A, WWTP B, and WWTP C) from Southeast, Queensland, Australia, from 26/05/2025 to 25/06/2025 with population ranging from 230,000 to 580,000. Simultaneously, two sets of Torpedo-style passive samplers (Schang et al. 2021), one for the same 24 h period, and the other for 1 week (~168 h), were deployed in the WWTP influent. The longer of these deployments was included to potentially enhance detection of low-to-moderate-prevalence or intermittently shed MeV across the study period (Chen and Bibby 2005). The sampling schedule is detailed in Supplementary Table ST1. Notably, there were no reported cases of MeV in the study catchments or the surrounding region in the four weeks prior to the commencement of this pilot study indicating a low incidence setting. One MeV case was notified in Brisbane in June 2025 during the sampling period.

### 2.2 influent Composite Wastewater Sample Concentration

Influent 24-hour composite wastewater samples were processed within 2 hours from arrival at the CSIRO environment laboratory (i.e., the primary laboratory, Lab A). An adsorption-extraction (AE) concentration method was used to concentrate viruses from influent wastewater samples (Akter et al., 2024). In the AE workflow, MgCl_2_ (Sigma-Aldrich, St. Louis, Missouri, USA) was added to each 50 mL wastewater sample to achieve a final concentration of 25 mM MgCl_2_. The remaining ~150 mL of wastewater sample was archived at −20°C for retrospective analysis or further processing to obtain additional nucleic acid if required. Each wastewater sample was filtered through 0.45 μm pore-size, putatively negatively charged MCE membrane (47-mm diameter, HAWP04700, Darmstadt, Germany) using a magnetic filter funnel (Pall Corporation, Port Washington, New York, USA) and a filter flask (Merck Millipore Ltd.). After filtration, the membrane was aseptically removed from the filter funnel using sterilised tweezers, rolled, and inserted into a 7-mL bead-beating tube (Axygen, CA, USA). Each bead-beating tube contained approximately 1 g of zirconium oxide beads (0.5 mm and 1 mm diameter; 1:1 w/w; Next Advance, Inc., Troy, NY, USA) for nucleic acid extraction. Each bead-beating tube containing the membrane (sample) was seeded with a known number (6,100 ± 400 GC) of murine hepatitis virus (MHV) to serve as an extraction control, with the objective of determining the MHV extraction recovery for each wastewater sample.

### 2.3 Torpedo Membrane Processing

The Torpedo-style passive samplers contain sorbent materials that sample virus particles and RNA from the water during the deployment period. Each Torpedo sampler contained two negatively charged gridded MCE membranes (Millipore, 0.45-μm), and a gauze, with one membrane from each sampler selected for nucleic acid extraction (the second stored as backup). The membrane was removed from the sampler using sterilized tweezers, rolled, and inserted into a 7-mL bead-beating tube (Axygen). Each bead-beating tube contained approximately 1 g of zirconium oxide beads (0.5 mm and 1 mm diameter; 1:1 w/w; Next Advance, Inc.) for nucleic acid extraction. Each bead-beating tube containing the membrane (sample) was seeded with known number (6,100 ± 400 GC) of MHV to serve as an extraction control. The remaining membranes and gauze were stored at −80°C for retrospective or further molecular analysis, if required.

### 2.4 Nucleic Acid Extraction

Nucleic acid extraction from each of the influent composite wastewater and the Torpedo membrane was performed using the RNeasy PowerWater Kit (Cat. No. 14700–50-NF, Qiagen). To each bead-beating tube containing the membrane and MHV, 850 μL of lysis buffer PM1 (Qiagen, Hilden, Germany), 150 μL of TRIzol reagent (Ambion, Thermo Fisher Scientific, Waltham, MA, USA), and 10 μL of β-mercaptoethanol (Cat. No. M6250-10ML, Sigma-Aldrich, St. Louis, MO, USA) were added. The tube contents were homogenized using a Precellys 24 tissue homogenizer (Bertin Technologies, Montigny-le-Bretonneux, France) at 9,000 rpm for three 15 s cycles, with a 10 s interval between cycles. After homogenization, the tubes were centrifuged at 4,000 *g* for 5 min to pellet filter debris, solids, and beads. DNase I solution was omitted from the protocol to enable simultaneous isolation of both DNA and RNA targets, supporting multi-pathogen detection. The sample was then eluted with 150 μL of nuclease-free water and stored at −20°C prior to qPCR/RT-PCR analysis. Concentrations and quality (*A*_260/280_ and *A*_260/230_) of nucleic acid samples were measured using a DeNovix DS-11 Series Spectrophotometer/Fluorometer (Wilmington, DE, USA).

### 2.5 RT-qPCR Assays

The quantification of MHV (Besselsen et al. 2022), MeV (Hummel et al. 2006) N gene and MeV M gene (Wu et al. 2024) was performed in Lab A using published RT-qPCR assays. The N gene assay detects B3, D4, D8, D9, H1 (MeV wild-type strains) and A (MeVV strains), whereas the M gene assay specifically detects the genotype A (MeVV strains). To independently confirm the results obtained in Lab A. RNA extracts were shared with an independent laboratory (PathWest laboratory; Western Australia; Lab B), where RT-PCR analysis of the N gene assay was performed. The N gene assay used by Lab B included bacteriophage MS2 as an internal process control (Chidlow et al. 2010; Tran et al. 2018). This assay detects MeV wildtype (B2, B3, D4, D8, D9, D11, G3 and H1) and MeVV strains (A).

The RT-qPCR analyses of MHV in Lab A were conducted in 20 μL reactions using TaqMan™ Fast Virus 1-Step Master Mix (Applied Biosystems, California, USA), with mixtures contained 5 μL of Supermix, 300 nM of each forward and reverse primer, 300 nM of probe, and 3 μL of nucleic acid. The RT-PCR analyses of MeV N gene in Lab A were performed in 20 μL reactions using TaqMan™ Fast Virus 1-Step Master Mix (Applied Biosystem, California, USA), with mixtures containing 5 μL of Supermix, 600 nM of each forward and reverse primer, 500 nM of probe, and 5 μL of nucleic acid. In lab B, RT-PCR analyses of MeV N gene were performed in 20 μL reactions using TaqMan™ Fast Virus 1-Step Master Mix (Applied Biosystem, California, USA), with mixtures containing 5 μL of Supermix, 400 nM of each forward and reverse primer, 200 nM of probe, and 5 μL of nucleic acid. The RT-PCR analyses of MeV M gene were performed (only in Lab A) in 20 μL reactions using TaqMan™ Fast Virus 1-Step Master Mix (Applied Biosystem, California, USA), with mixtures containing 5 μL of Supermix, 900 nM of each forward and reverse primer, 250 nM of probe, and 5 μL of nucleic acid. The primer and probe sequences, their concentrations in reactions, and cycling parameters are detailed in Table 1. Gamma-irradiated viruses and gBlocks were used for standard curves and positive controls. In Lab A, all RT-qPCR reactions were performed in triplicate on a Bio-Rad CFX96 thermal cycler (Bio-Rad Laboratories, Hercules, California, USA) with automated settings for threshold and baseline, alongside PCR positive/standard and negative controls. In Lab B, RT-qPCR reactions were performed in duplicate on a CFX96 Touch Real-Time PCR Detection System (Bio-Rad Laboratories, Hercules, California, USA), alongside PCR positive/standard and negative controls.

**Table 1:**
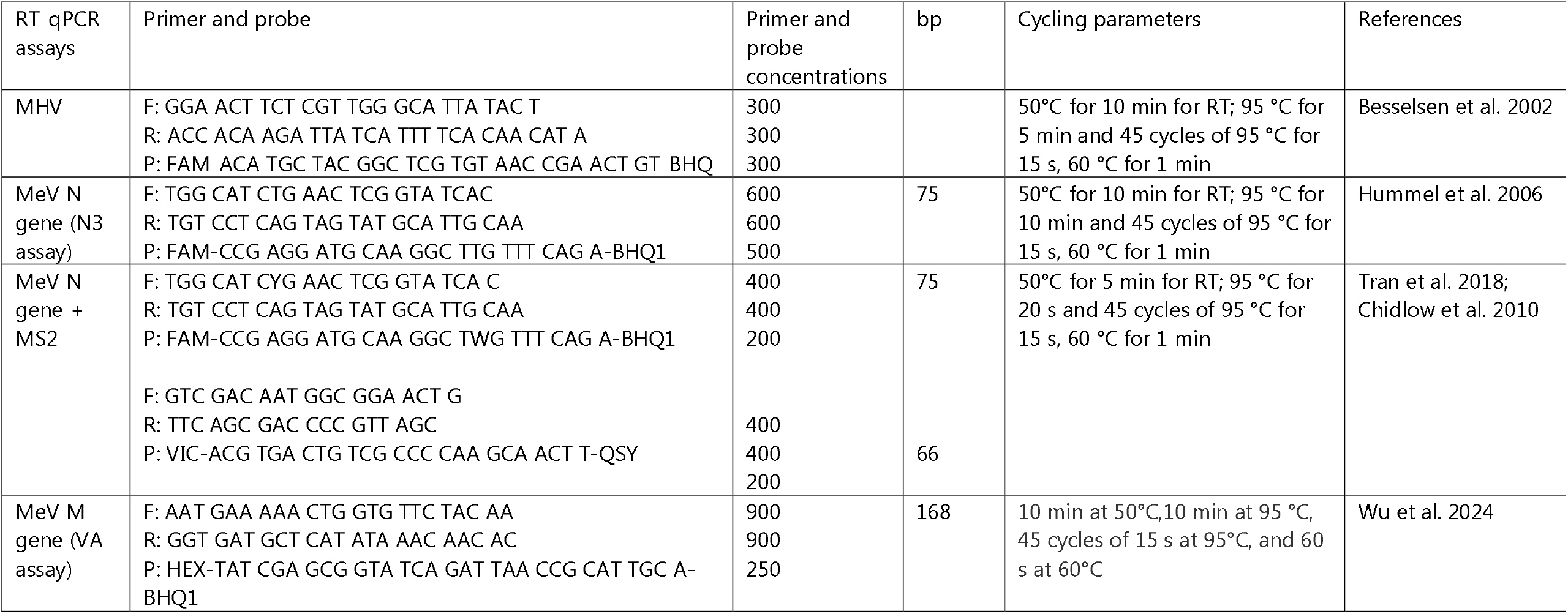
Primer and probes used in this study.

### 2.6 Sequence confirmation

Two methods were trialed for sequence confirmation: Matrix gene sequencing and Tiled-amplicon sequencing (performed in Lab B). The Matrix gene sequencing used a previously described nested PCR assay targeting a 187 bp region of the matrix (M) gene (Jin et al. 1996) with minor primer modifications. This assay has been routinely used in Lab B for genotyping for clinical and wastewater samples, with historical concordance to N450 sequencing assays performed at reference laboratories. Duplicate first-round PCR reactions were prepared in a total volume of 20 µL containing 5 µL of nucleic acid extract, TaqMan™ Fast Virus 1-Step Multiplex Master Mix for qPCR (No ROX), and primers (F: TCA GAG TCA TAG ATC CTG GT; R: AAC AAC TAT GTC AAR CTC AG) at a final concentration of 500 nM. Each first-round product was used as template for the second-round PCR, prepared in a total volume of 20.5 µL containing 0.5 µL of first-round product, 1× PE Buffer II, 2 mM MgCl_2_, 0.2 mM of each dNTP, 0.5 U PE TaqGold, 0.01% BSA, and 200 nM of each primer (F: ATA GAT CCT GGT CTA GGC; R: ACT ATG TCA WGC TCA GTG). Second-round PCR products were electrophoresed on an agarose gel and examined for the expected amplicon size.

The Tiled-amplicon sequence confirmation used a previously published tiled-amplicon whole-genome sequencing assay (Kent et al. 2024; <u>GitHub - artic-network/artic-measles: Repository for realtime sequencing of measles virus using the MinION</u>) with minor in-house primer modifications with approximately 400 bp amplicon sizes spanning the measles virus genome. This assay has been implemented in Lab B last year and is used for cluster analysis of clinical MeV sequences. Additionally, it is used on Western Australian wastewater samples successfully generating partial genomes with sufficient viral concentration. Duplicate cDNA synthesis reactions were prepared in a total reaction volume of 20 µL containing 5 µL of nucleic acid extract and LunaScript® RT SuperMix Kit.

Duplicate cDNA synthesis reactions were prepared in a total volume of 20 µL containing 5 µL of nucleic acid extract and the LunaScript® RT SuperMix Kit. The synthesised cDNA was then used as template for PCR amplification in two separate non-overlapping tiled-amplicon pools. Each pool was prepared in a total reaction volume of 25 µL containing 6 µL of cDNA, Q5® Hot Start High-Fidelity 2× Master Mix, and 0.015 µM of each primer. PCR products from both pools were combined in equal volumes and purified using a 1× AMPure SPRI bead ratio. Libraries were prepared using the Illumina Nextera XT DNA Library Preparation Kit following the manufacturer’s protocol, with reagent volumes reduced by half and unique dual indexing applied. Sequencing was performed on the Illumina MiniSeq platform. Run quality control and data analysis were conducted using a custom in-house bioinformatics pipeline.

### 2.7 Extraction Recovery

Following nucleic acid extraction (as described in Section 2.4), the MHV GC number in each sample was determined using the RT-qPCR assay outlined in Section 2.5. The MHV extraction recovery was calculated using the formula:

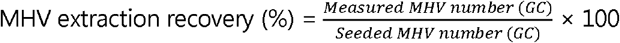

The initial MHV number was based on the seeded amount (6,100 ± 400 GC), and the measured number was derived from the RT-qPCR quantification. All recovery calculations were performed in RT-qPCR triplicate, with results averaged to account for variability.

### 2.8 QA/QC and Data Interpretation

Nucleic acid extraction and PCR setup in Lab A were performed in separate rooms to minimize potential contamination. The amplification efficiencies (E), correlation coefficient (R^2^), and y-intercepts for calibration curves were assessed based on the Minimum Information of Publication of Quantitative Real-Time PCR Experiments guidelines (Bustin et al. 2009). An increase in Cq >2 cycles indicated potential inhibition, with higher values suggesting stronger inhibition effects (Ahmed et al. 2020). Wastewater samples were classified as positive if amplification was detected in at least one out of three replicates within 40 cycles. Samples were considered indeterminate when the Cq value was between 40 to 45. A sample was considered not-detected (ND) or negative when none of the three replicates showed amplification. A sample was considered quantifiable when all three replicated amplified.

## 3. Results and Discussion

PCR inhibition was observed in six of 33 wastewater nucleic acid samples in Lab A, with three samples from each of the composite and passive sampling method displaying inhibition. These inhibited samples were diluted tenfold and reanalyzed, successfully mitigating the effect, while RT-qPCR negative controls showed no detectable amplification (ND), confirming the absence of cross-contamination. This inhibition rate (18.2%) highlights matrix-related challenges in WWS, though the dilution approach proved effective, aligning with previous studies on virus detection in wastewater (Ciesielski et al. 2021; Ozawa et al. 2025).

Extraction recovery, assessed using MHV as a process control, varied markedly across sampling methods and wastewater treatment plants (WWTPs). Pooled data across all WWTPs over four weeks revealed mean extraction efficiencies of 49.2 ± 14.1% for 24-h passive samples (n = 11), 33.3 ± 15.8% for 168-h passive samples (*n* = 12), and 27.3 ± 7.3% for 24-h composite samples (*n* = 12) (Fig. 1). The highest recovery (70.5%) was observed in a 24-h passive sample collected at WWTP A on April 4–5, 2025, while the lowest (11.5%) was recorded in a 168-h passive sample from WWTP C on June 2–9, 2025 (Supplementary Table ST2). These results demonstrate that the 24-h passive sampling method consistently yielded the highest average recovery, while the 24-h composite samples showed the lowest recovery.

**Fig. 1.**
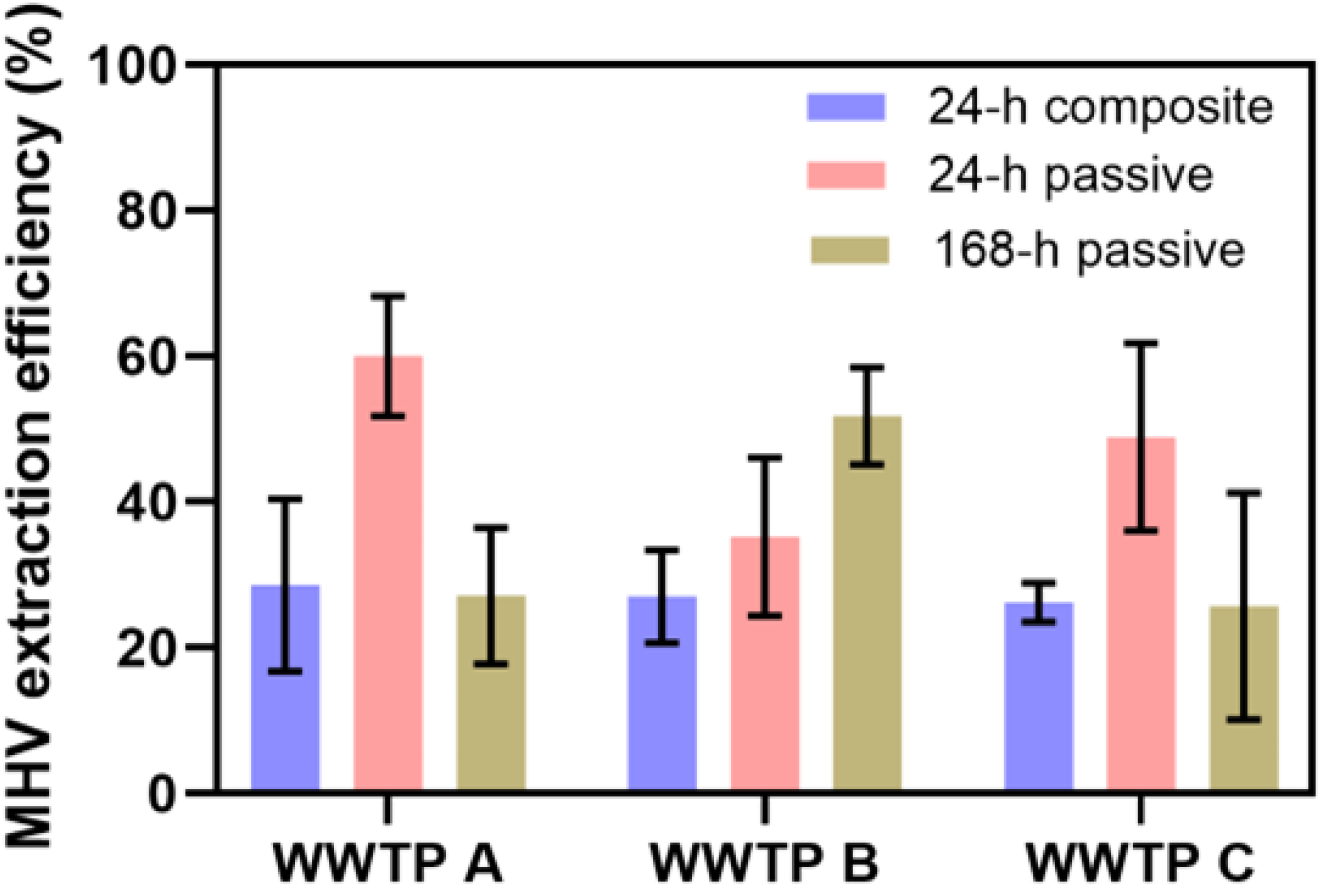
Nucleic acid extraction recovery (%) of Murine Hepatitis Virus (MHV) for pooled 24-h composite wastewater, 24-h passive, and 168-h passive membrane samples across three WWTPs.

The observed variability in extraction recovery (mean 27.3 – 49.2% by sample type, and range of 11.5–70.5%) underscores method/sample-specific performance differences and highlights the need for optimization to ensure consistent sensitivity across sampling types. Notably, the extraction recovery efficiencies reported here are higher than those observed in a previous study by Habtewold et al. (2022), which reported extraction recovery rates ranging from 1.6% to 24% using phi6-seeded composite wastewater and Torpedo membranes. In our study, recovery was assessed specifically for the nucleic acid extraction step, as the passive sampling method did not require a concentration step.

Despite equal MHV seeding across bead beating tubes, differences in extraction recovery suggest that sample composition and physical characteristics significantly influenced extraction outcomes. The higher extraction recovery in 24-h passive samples may be attributed to lower levels of suspended solids and particulate matter, which can interfere with extraction by clogging spin columns and reducing nucleic acid yield. This observation aligns with findings from Habtewold et al. (2022), who also reported higher recovery from passive membranes.

In contrast, membranes from the 168-h passive and 24-h composite samples contained visibly more solids and excess liquid in the bead-beating tubes. During lysate transfer following bead beating, viral targets may have remained bound to bead particles and retained in the bead-beating tube, contributing to reduced recovery. These findings highlight the critical impact of sample matrix effects on extraction recovery and underscore the importance of method- and site-specific evaluations to ensure accurate quantification and reliable trend analysis in WWS.

Wastewater samples from three sites, collected between May 28, 2025 and June 23, 2025, yielded two notable MeV RNA detections at WWTP B on May 26–27, 2025, from 24-h composite and 24-h Torpedo samples collected in parallel (Table 2). No MeV was detected on the 1-week passive sample collected May 26 – June 2 at the same site. This catchment, which has an estimated 584,000 persons (2021 census), showed MeV RT-PCR in 3 of 3 replicates for the composite sample (Cq = 37.4, 38.3, 36.7), and 2 of 3 replicates for the passive sample (Cq = 38.9, ND, 37.7). The contemporaneous detection across two sampling methods (composite and passive) strengthens the result’s reliability despite high Cq values indicating low viral load. Notably, no clinical measles cases were reported in Brisbane during May (https://nindss.health.gov.au/pbi-dashboard/), making this an unexpected detection. One MeV case was notified in Brisbane in June 2025 during the study period, and although epidemiological information available to the study did not indicate an exposure window consistent with late May, it remains theoretically possible that prodromal□phase shedding could contribute to wastewater signals.

**Table 2:** Detection of Measles Virus (MeV) in wastewater samples from three WWTPs using different sampling approaches.

| WWTPs |  |  | MeV positive detection (Cq values) |  |  |  |
| --- | --- | --- | --- | --- | --- | --- |
|  | Sample type | Dates | CSIRO MeV N gene (N3) assay | PathWest MeV N gene assay | CSIRO MeV M gene assay | PathWest Whole genome and tiled amplicon sequencing |
| WWTP A | Composite 24-h | 28/05/2025 - 29/05/2025 | ND | NT | NT | NT |
|  | Passive 24-h | 28/05/2025 - 29/05/2024 | ND | NT | NT | NT |
|  | Passive 168-h | 28/05/2025 - 04/6/2025 | ND | NT | NT | NT |
|  | Composite 24-h | 04/06/2025 - 05/06/2025 | ND | NT | NT | NT |
|  | Passive 24-h | 04/06/2025 - 05/06/2025 | ND | NT | NT | NT |
|  | Passive 168-h | 04/06/2025 - 11/06/2025 | ND | NT | NT | NT |
|  | Composite 24-h | 11/06/2025 - 12/06/2025 | ND | NT | NT | NT |
|  | Passive 24-h | 11/06/2025 - 12/06/2025 | ND | NT | NT | NT |
|  | Passive 168-h | 11/06/2025 - 18/06/2025 | ND | NT | NT | NT |
|  | Composite 24-h | 18/06/2025 - 19/06/2025 | ND | NT | NT | NT |
|  | Passive 24-h | 18/06/2025 - 19/06/2025 | ND | NT | NT | NT |
|  | Passive 168-h | 18/06/2025 - 25/06/2025 | ND | NT | NT | NT |
| WWTP B | Composite 24-h | 26/5/2025 - 27/05/2025 | POS (37.4, 38.3, 36.7) | POS (39.2, 36.2) | POS (37.5, 38.1, 37.6) | ND |
|  | Passive 24-h | 26/5/2025 - 27/05/2025 | POS (38.9, ND, 37.7) | ND | NT | ND |
|  | Passive 168-h | 26/5/2025 to 02/06/2025 | ND | NT | NT | NT |
|  | Composite 24-h | 09/06/2025 - 10/06/2025 | ND | NT | NT | NT |
|  | Passive 24-h | 09/06/2025 - 10/06/2025 | ND | NT | NT | NT |
|  | Passive 168-h | 09/06/2025 - 16/06/2025 | ND | NT | NT | NT |
|  | Composite 24-h | 16/06/2025 - 17/06/2025 | ND | NT | NT | NT |
|  | Passive 24-h | 16/06/2025 - 17/06/2025 | ND | NT | NT | NT |
|  | Passive 168-h | 17/06/2025 - 23/06/2025 | ND | NT | NT | NT |
| WWTP C | Composite 24-h | 26/05/2025 - 27/05/2025 | ND | NT | NT | NT |
|  | Passive 24-h | 26/05/2025 - 27/05/2025 | ND | NT | NT | NT |
|  | Passive 168-h | 26/05/2025 - 02/06/2025 | ND | NT | NT | NT |
|  | Composite 24-h | 02/06/2025 - 03/06/2025 | ND | NT | NT | NT |
|  | Passive 24-h | 02/06/2025 - 03/06/2025 | ND | NT | NT | NT |
|  | Passive 168-h | 02/06/2025 - 09/06/2025 | ND | NT | NT | NT |
|  | Composite 24-h | 09/06/2025 - 10/06/2025 | ND | NT | NT | NT |
|  | Passive 24-h | 09/06/2025 - 10/06/2025 | ND | NT | NT | NT |
|  | Passive 168-h | 09/06/2025 - 16/06/2025 | ND | NT | NT | NT |
|  | Composite 24-h | 16/06/2025 - 17/06/2025 | ND | NT | NT | NT |
|  | Passive 24-h | 16/06/2025 - 17/06/2025 | ND | NT | NT | NT |
|  | Passive 168-h | 16/06/2025 - 23/06/2025 | ND | NT | NT | NT |

POS: Positive; ND: Not detected; NT: Not tested.

Lab B was able to amplify measles genetic fragments using N gene RT-qPCR assay (detects B3, D4, D8, D9, H1 (MeV wild-type strains) and A (MeVV strains) in 24-h composite sample. Lab B then attempted sequencing using two methods to confirm the genotype, with a low expectation of success given the high Cq values. The Matrix gene sequencing primers did not amplify any PCR products, therefore, sequencing was not conducted. The Tiled amplicon sequencing also did not map any reads to MeV reference genomes. Later, a specific RT-PCR assay (MeV M gene assay by Wu et al. 2024) for MeVV used by CSIRO laboratory yielded positive detection indicating likely vaccine-related shedding by an individual(s) in 3 of 3 replicates tested from the composite sample. This aligns with Canadian reports of post-immunization detections (Tomalty et al. 2025), though such report remains rare globally. However, given the RT-qPCR assays used in this study (the N gene assay detects both MeV wild-type and MeVV strains, whereas M gene assay detects only MeVV strains), the possibility that wild-type strains were also present could not be ruled out.

This finding has two key implications. First, it suggests at least one case of vaccine-derived measles shedding in the study catchment in the absence of reported clinical cases (except one case in June), with an international airport’s inclusion within the wastewater catchment raising the possibility of a travel-related source, a critical consideration for border health surveillance. Second, the rarity of MeV detection in WWS, even from MeVV strains, underscores the method’s sensitivity and the need for genotypic differentiation to distinguish MeV wild-type from MeVV and inform appropriate local public health action (Chen and Bibby 2025; Gan et al. 2025; Paulos et al. 2025). The MeV N gene assay used in this study detects both MEV wild-type and MeVV strains; therefore, incorporating assays capable of detecting these strains separately will be valuable for accurate data interpretation and targeted public health response (Wu et al. 2024).

The sparse detection (2/33 samples, 6.1% from a WWTP) of MeVV strain RNA suggest the potential for WWS of MeV. The findings from this study (with samples collected 26/05/2025 - 25/06/2025) take on added significance considering Queensland’s 2025 measles outbreaks, with 30 confirmed cases reported by 12 November 2025, many of which were imported cases among people returning or travelled from overseas (https://www.health.qld.gov.au/newsroom/doh-media-releases/public-health-alert-measles; https://bsphn.org.au/news/public-health-alert-measles-november-2025). Notable exposure periods occurred from April 7–9 in Brisbane (https://metronorth.health.qld.gov.au/news/measles-alert-for-locations-in-brisbane), August 19–23 in the West Moreton region (https://www.westmoreton.health.qld.gov.au/about-us/news/measles-alert-for-the-west-moreton-region), September 19–21 in Brisbane and Mooloolaba (https://metronorth.health.qld.gov.au/news/measles-alert-brisbane-sunshine-coast), and October 9–13 in Caloundra on the Sunshine Coast (https://www.sunshinecoast.health.qld.gov.au/about-us/news/articles/measles-alert-october-2025) (all within 150 km of Brisbane).

Wastewater samples from these catchments were not tested during these periods. WWS of MeV may provide the capabilities to detect MeV signals early/earlier than clinical surveillance to inform public health decision making. This underscores the value of sustained WWS as a sensitive tool for monitoring highly contagious pathogens like MeV in vaccinated and unvaccinated populations. It may be particularly useful in settings with elevated risk of measles importations such as airports where early signals can complement clinical surveillance and enhance outbreak detection and decision-making. Future studies should broaden its scope to encompass different seasons and refine detection techniques to better attribute outbreak sources.

Vaccination timing data for the sampled catchments was not accessible; therefore, it was not possible to determine whether the detections coincided with intensified vaccination campaigns. However, given the low detection frequency and the short duration of MeVV strains shedding typically observed post-immunization, these detections were more likely incidental findings associated with routine vaccination rather than systematic shedding linked to outbreak-driven immunization efforts. Additional investigations are warranted to elucidate the influence of genotype and vaccination status on shedding dynamics and their implications for the effectiveness of WWS for MeV.

In conclusion, this pilot study demonstrates the feasibility of using WWS for monitoring MeV in Australia, with the detection in wastewater offering a proof-of-concept for early signal detection. Integrating genotypic analysis and advanced methods could elevate the role of WWS for national preparedness and response, including for vaccine preventable, high-R_0_ pathogens like measles. This study also provides the first evidence that passive sampling can be successfully applied for MeV detection in wastewater, offering a practical, low-maintenance approach that enhances sensitivity for identifying both vaccine-derived shedding and potentially wild-type MeV. Although not applied in this study, the incorporation of a wild-type–specific assay will strengthen the ability to distinguish circulating MeV wild-type virus from MeVV related signals. Furthermore, the successful detection of MeV using passive sampling in this study highlights its potential for deployment in diverse contexts, including non-sewered settings where composite sampling is not feasible, thereby expanding the reach and adaptability of wastewater surveillance.

## Data Availability

All data produced are available in the manuscript

## Acknowledgement

The authors acknowledge funding support from the Australian Department of Health and Aged Care (now the Department of Health, Disability and Ageing) and the interim Australian Centre for Disease Control (now Australian Centre for Disease Control) (Project ID: E25-87539), and advice from The Queensland Department of Health to include measles among emerging pathogens tested in Queensland. This work was undertaken by Promoting Health4All Pty Ltd (lead contractor) in collaboration with CSIRO (the primary laboratory) and PathWest (an independent laboratory) to conduct time-limited WWS trials aimed at developing and enhancing sampling and analytical methods for emerging public health threats such as MeV that remain at an early stage of methodological and field application. Our sincere thanks to both the Water Unit and Public Health Intelligence Branch of Queensland Health for their support throughout the Wastewater Surveillance - Preparedness for Emerging Priority Pathogens (WS-PEPP) Project.

