## Supplementary Tables for "Detection of Measles Virus RNA in Wastewater: Monitoring for Wild-Type and Vaccine-Derived Strains in a National Preparedness Trial"

**Supplementary Table ST1**

Sampling schedule for composite and torpedo passive samplers at three wastewater treatment plants (WWTPs)

| WWTPs | Sample Type | Week 1 (in) | Week 1 (out) | Week 2 (in) | Week 2 (out) | Week 3 (in) | Week 3 (out) | Week 4 (in) | Week 4 (out) |
| --- | --- | --- | --- | --- | --- | --- | --- | --- | --- |
| WWTP A | 24-h Composite | 28/05/2025 | 29/05/2025 | 04/06/2025 | 05/06/2025 | 11/06/2025 | 12/06/2025 | 18/06/2025 | 19/06/2025 |
|  | 24-h passive | 28/05/2025 | 29/05/2025 | 04/06/2025 | 05/06/2025 | 11/06/2025 | 12/06/2025 | 18/06/2025 | 19/06/2025 |
|  | 168-h Passive | 28/05/2025 | 04/06/2025 | 04/06/2025 | 11/06/2025 | 11/06/2025 | 18/06/2025 | 18/06/2025 | 25/06/2025 |
| WWTP B | 24-h Composite | 26/05/2025 | 27/05/2025 | 02/06/2025 | 03/06/2025 | 09/06/2025 | 10/06/2025 | 16/06/2025 | 17/06/2025 |
|  | 24-h passive | 26/05/2025 | 27/05/2025 | 02/06/2025 | 03/06/2025 | 09/06/2025 | 10/06/2025 | 16/06/2025 | 17/06/2025 |
|  | 168-h Passive | 26/05/2025 | 02/06/2025 | 02/06/2025 | 09/06/2025 | 09/06/2025 | 16/06/2025 | 16/06/2025 | 23/06/2025 |
| WWTP C | 24-h Composite | 26/05/2025 | 27/05/2025 | 02/06/2025 | 03/06/2025 | 09/06/2025 | 10/06/2025 | 16/06/2025 | 17/06/2025 |
|  | 24-h passive | 26/05/2025 | 27/05/2025 | 02/06/2025 | 03/06/2025 | 09/06/2025 | 10/06/2025 | 16/06/2025 | 17/06/2025 |
|  | 168-h Passive | 26/05/2025 | 02/06/2025 | 02/06/2025 | 09/06/2025 | 09/06/2025 | 16/06/2025 | 16/06/2025 | 23/06/2025 |

**Supplementary Table ST2**

MHV extraction recovery (%) for composite and passive wastewater samples across three WWTPs

| WWTPs | Sample type | Dates | Extraction recovery (%) |
| --- | --- | --- | --- |
| WWTP A | Composite 24-h | 28/05/2025 - 29/05/2025 | 44.0 ± 0.00 |
|  | Passive 24-h | 28/05/2025 - 29/05/2024 | 58.4 ± 0.00 |
|  | Passive 168-h | 28/05/2025 - 04/06/2025 | 38.4 ± 4.52 |
|  | Composite 24-h | 04/06/2025 - 05/06/2025 | 15.3 ± 2.16 |
|  | Passive 24-h | 04/06/2025 - 05/06/2025 | 70.5 ± 2.85 |
|  | Passive 168-h | 04/06/2025 - 11/06/2025 | 27.5 ± 2.19 |
|  | Composite 24-h | 11/06/2025 - 12/06/2025 | 27.5 ± 1.13 |
|  | Passive 24-h | 11/06/2025 - 12/06/2025 | 60.3 ± 19.8 |
|  | Passive 168-h | 11/06/2025 - 18/06/2025 | 15.5 ± 3.50 |
|  | Composite 24-h | 18/06/2025 - 19/06/2025 | 27.5 ± 1.53 |
|  | Passive 24-h | 18/06/2025 - 19/06/2025 | 50.7 ± 0.00 |
|  | Passive 168-h | 18/06/2025 - 25/06/2025 | 26.8 ± 0.00 |
| WWTP B | Composite 24-h | 26/05/2025 - 27/05/2025 | 19.8 ± 1.65 |
|  | Passive 24-h | 26/05/2025 - 27/05/2025 | 35.0 ± 5.30 |
|  | Passive 168-h | 26/05/2025 - 02/06/2025 | 58.4 ± 0.00 |
|  | Composite 24-h | 09/06/2025 - 10/06/2025 | 29.6 ± 3.24 |
|  | Passive 24-h | 09/06/2025 - 10/06/2025 | 24.4 ± 0.98 |
|  | Passive 168-h | 09/06/2025 - 16/06/2025 | 45.1 ± 1.86 |
|  | Composite 24-h | 16/06/2025 - 17/06/2025 | 31.6 ± 1.31 |
|  | Passive 24-h | 16/06/2025 - 17/06/2025 | 46.1 ± 1.86 |
|  | Passive 168-h | 17/06/2025 - 23/06/2025 | 52.0 ± 4.14 |
| WWTP C | Composite 24-h | 26/05/2025 - 27/05/2025 | 28.1 ± 1.13 |
|  | Passive 24-h | 26/05/2025 - 27/05/2025 | 46.1 ± 1.86 |
|  | Passive 168-h | 26/05/2025 - 02/06/2025 | 47.3 ± 3.34 |
|  | Composite 24-h | 02/06/2025 - 03/06/2025 | 24.5 ± 2.60 |
|  | Passive 24-h | 02/06/2025 - 03/06/2025 | 51.9 ± 2.14 |
|  | Passive 168-h | 02/06/2025 - 09/06/2025 | 11.5 ± 0.00 |
|  | Composite 24-h | 09/06/2025 - 10/06/2025 | 23.3 ± 0.00 |
|  | Passive 24-h | 09/06/2025 - 10/06/2025 | 33.2 ± 2.35 |
|  | Passive 168-h | 09/06/2025 - 16/06/2025 | 25.7 ± 2.82 |
|  | Composite 24-h | 16/06/2025 - 17/06/2025 | 28.8 ± 2.04 |
|  | Passive 24-h | 16/06/2025 - 17/06/2025 | 64.3 ± 5.12 |
|  | Passive 168-h | 16/06/2025 - 23/06/2025 | 18.2 ± 3.27 |
